# Reconsolidation blockade with propranolol as a novel treatment for chronic low-back pain: a double-blind randomized placebo-controlled feasibility study

**DOI:** 10.1101/2025.09.12.25335577

**Authors:** Alexia Coulombe-Leveque, Sylvie Lafrenaye, Alain Brunet, Serge Marchand, Guillaume Léonard

## Abstract

**Purpose:** Nociplastic pain is often characterized by maladaptive plasticity in the nervous system similar to that observed in patients with post-traumatic stress disorder (PTSD). The aim of this study was to investigate whether reconsolidation therapy, a treatment for PTSD consisting in reactivating (through trauma narrative) the synapses encoding the excessive threat response and blocking their reconsolidation using propranolol, is feasible in patients with nociplastic low-back pain.

**Patients and methods: Design:** triple-blind, placebo-controlled feasibility study.

Population: 24 adults with chronic (>6 months) nociplastic low-back pain with no comorbid PTSD or contra-indication to propranolol.

Intervention: Pain education (10 short videos) and 6 weekly sessions of reconsolidation therapy with propranolol (n=12) or placebo (n=12) administered orally 1h pre-reactivation).

Outcome measures:

Feasibility: recruitment rates, adverse events (frequency/severity).

Effect of intervention: Brain Pain Inventory (BPI) and other self-reported pain questionnaires, 4 weeks post-intervention.

**Results:** Sixty-six patients were screened over 6 months; 24 participants were enrolled; 2 dropped out. Adverse events were mild and infrequent (asymptomatic decrease in heart rate (n=4), headache and nausea (n=1). No clinically meaningful difference was observed between the two groups on the pain questionnaires. However, we noted prevalent catastrophic/kinesiophobic discourse during the sessions, and the reactivation methods appeared to have been suboptimal for the population.

**Conclusion:** Reconsolidation therapy is a feasible intervention for chronic pain. Preliminary results suggest no effect on pain symptoms. Additional studies are warranted to assess the adequacy of reactivation procedures (proper reactivation being required to trigger reconsolidation), and to investigate whether the absence of negative pain beliefs might be a prerequisite (unmet in this study) for the success of the intervention.

## Introduction

Chronic nociplastic pain is characterized by plastic changes within the central nervous system^1^, notably by a shift in pain representation within the brain from the somatosensory areas towards the limbic system ^2,3^.

The amygdala, a key component of the limbic system involved in threat learning, has emerged as a particular region of interest in the physiopathology of nociplastic pain. Evidence suggests that the amygdala plays a pronociceptive role in the context of chronic pain ^4–9^, and morphological as well as functional alterations have been observed in the amygdala of patients with nociplastic pain ^9–17^. In parallel, animal studies have shown that chemogenetic activation of the amygdala can induce or heighten pain behaviours, while its silencing inhibits or prevents them (see ^6,11^ for reviews).

These changes in the limbic system are reminiscent of those observed in patients with post-traumatic stress disorder (PTSD) ^9,18–20^, who exhibit increased amygdala reactivity after excessive threat learning ^21–23^. A new treatment for PTSD was developed in the 2010s, which specifically aims to reverse these plastic changes in the amygdala: reconsolidation therapy (RT) ^24,25^. RT is based on synaptic reconsolidation, a process rediscovered by Susan Sara and Jean Przybyslawski in the late ’90s and popularized by Karim Nader in 2000 ^26–28^. Its principles are rooted in the concept that a consolidated synaptic trace, when reactivated under certain conditions, requires *re*-consolidation; during this period, the synapse becomes temporarily unstable and malleable, and certain molecules can be administered to interfere with its reconsolidation, which results in the long-term weakening of the synaptic connection ^29,30^. Such agents include protein-synthesis inhibitors and, for reconsolidation blockade in the amygdala, the lipophilic beta-blocker propranolol ^26,31,32^. RT for the treatment of PTSD consists of two elements: i) a ‘reactivation’, wherein patients are asked, for instance, to write an autobiographical script of their traumatic experience (so as to cause the underlying synaptic connections to undergo reconsolidation); and ii) administration of propranolol (to disrupt the reconsolidation of the reactivated synaptic traces in the amygdala, thereby weakening them and normalizing the pathologically heightened threat response) ^25,33^.

Given its aim to reverse maladaptive amygdala plasticity – and the evidence implicating such plasticity in nociplastic pain – RT represents a promising therapeutic candidate warranting investigation. Yet, to date and to the best of our knowledge, no clinical study has investigated the feasibility and/or potential benefits of RT in patients with chronic pain.

The primary objective of this study was to assess the feasibility and acceptability of a modified RT intervention (6 weekly sessions of RT with placebo or propranolol) delivered to adults with chronic nociplastic low back pain. The secondary objective was to gather preliminary data on the effect of the intervention on patient-reported pain symptoms (physical function, pain intensity, emotional function).

## Material and methods

### Study Design

We conducted a double-blind, randomized, placebo-controlled feasibility study. A feasibility design was selected because reconsolidation therapy had never been studied as a possible treatment for chronic pain and requires the off-label use of propranolol, which raised important considerations in terms of recruitment (rates and selection criteria) and safety. The research protocol was approved by the local institutional ethics board (Comité d’éthique de la recherche du CIUSSS de l’Estrie-CHUS) and a non-objection letter was obtained from Health Canada, Division 5 on August 27^th^, 2021. The study was registered in Clinical Trials (NCT05085782) on September 27^th^, 2021. The study took place at the Research Center of the CIUSSS-de-l’Estrie-CHUS (Sherbrooke, Québec, Canada). Signed informed consent was obtained for all participants. There were no deviations from the research protocol during the trial.

### Monitoring

An independent study monitor conducted a full review of the study documentation (eg, operations manual, delegation log, equipment log, medication handling and storage procedures, contingency plan, ethical training certificates, etc.), as well as two data handling quality assessments.

### Setting

The study took place at the Research Center of the CIUSSS-de-l’Estrie-CHUS (Sherbrooke, Québec, Canada) from February to October 2022.

### Participants

This study was conducted in adults with chronic nociplastic low-back pain. (The study protocol was written before the term “nociplastic pain” gained widespread recognition and acceptance – as such, it uses the term “central sensitization” as operationalized by Nijs ^34^. However, seeing as this operationalization is consistent with the concept of “nociplastic pain” as defined by the IASP ^1^, this manuscript will use the term “nociplastic pain” to reflect the latest science.) Inclusion criteria were: 1) age 18 to 65 years old; 2) ability to understand spoken and written French; 3) low-back pain present for more than 6 months; 4) average daily pain intensity greater than 3/10 (verbal numeric scale); and 5) presence of nociplastic pain. This last criterion was assessed using the algorithm developed by Nijs et al. ^34^, which states that the presence of central sensitization can be suspected if neuropathic pain is unable to explain the clinical presentation, and if patients present 1) pain of disproportionate intensity relative to the nature and severity of the lesion ; and 2a) diffuse or neurologically implausible pain distribution ; or 2b) score of 40 or higher on the Central Sensitization Inventory (CSI), part A ^35^. These were assessed via a full standardized clinical evaluation (performed by ACL, physiotherapist).

Exclusion criteria were: 1) contraindication to propranolol; 2) contraindication to reconsolidation therapy ^33^ (eg, acute suicidal ideation; substance abuse); 3) comorbid PTSD; 4) recent (<3 years) lumbar surgery; 5) recent (<3 months) change in treatment; 6) involvement in a lawsuit related to the pain condition.

### Recruitment

Participants were recruited between February and August 2022 through adverts on social media and posters in medical clinics, physiotherapy clinics, and at the local hospital (convenience sampling). Eligibility was assessed through a preliminary phone screening and an in-person admissibility visit.

### Intervention

The intervention consisted of pain education and 6 weekly sessions of reconsolidation therapy with propranolol or a placebo.

### Pain Education

Prior to the first intervention session, participants watched 10 educational videos on pain neuroscience (2–4 minutes each). These videos were produced for the study by an experienced physiotherapist currently practicing with a chronic pain population (ACL), in collaboration with two patient-partners. Pain education was included in both groups as it is a recommended treatment for chronic pain treatment ^36^, thereby making the intervention multimodal and ensuring that the experimental treatment was not compared to a placebo alone.

### Reconsolidation Therapy

The experimental intervention included 6 weekly sessions of reconsolidation therapy ^25,33^, which combines a brief (10-15 mins) reactivation procedure with administration of propranolol (or a placebo). The reactivation procedure, which for PTSD revolves around a narrative script of the traumatic event, was modified for the chronic pain population by a transdisciplinary team including experts in chronic pain, experts in reconsolidation therapy, and a patient partner. Three distinct reactivation themes were developed (to increase mismatch between sessions ^37^) and used sequentially during each of the six intervention sessions (Theme A: Sessions 1 and 4; Theme B: Sessions 2 and 5; and so on).

Theme A, dubbed “Painful Event”, followed the PTSD treatment protocol most closely: patients were asked to verbally recount (out loud) their *most salient* episode of low back pain, in the first person, present tense, and to describe contextual and environment cues (eg sounds, odours, and other physical sensations associated with the event).

Theme B, dubbed “Nightmare Scenario”, was inspired by reconsolidation blockade as applied for the treatment of phobias ^38,39^ and from studies on Picture Imagination Tasks ^40^: participants were instructed to visualize a ‘nightmarish scenario’ related to their pain (eg having to perform particularly painful tasks or movements).

Theme C, dubbed “Negative Emotions”, was based on the well-established role of the amygdala in negative emotions, and the similarly well-established link between negative emotions and pain ^6,9,12,13,41,42^. Participants were asked to describe and try to feel the emotions they typically felt during a painful episode (low back-related).

### Propranolol

This study used short-acting propranolol (Teva-Canada) as the reconsolidation-blocking agent. The propranolol was encapsulated to look identical to the placebo (corn starch) by Gentès & Bolduc Pharmacists Canada. Dosage was the same as is used for PTSD ^25,33^: 40, 60 or 80 mg per session (based on height and self-reported biological sex, see Table 1). As per the PTSD treatment protocol, propranolol (or placebo) capsules were ingested at the beginning of each experimental session (along with a small snack to facilitate absorption ^43^) one hour prior to the reactivation procedure, so that the reactivation theoretically coincided with the peak plasma concentration of propranolol ^44^.

**Table 1.** Propranolol Dosage. Table 1 shows the dosage of propranolol (or placebo) based on height and (self-reported) biological sex.

|  | Women | Men |
| --- | --- | --- |
| 40 mg | <165 cm | <155 cm |
| 60 mg | 165-185 cm | 155-175 cm |
| 80 mg | >185 cm | >175 cm |

### Timeline of visits

**V0.** After an initial phone screening, prospective participants attended an admissibility visit (V0) at the research center. During this visit, a standardized clinical assessment was conducted by ACL to confirm eligibility, and participants completed the baseline pain questionnaires (see *outcome measures*).

**V1-V6**. Participants attended six weekly sessions of RT (V1 to V6).

At the beginning of each session, participants ingested the propranolol (or placebo) capsules and then engaged in a calming activity for 1h, such as reading or using a screen, while waiting for the medication to take effect ^44^. Heart rate and blood pressure were measured before medication intake and again 30 and 60 minutes later. After the 60-min vitals monitoring, the therapist performed a brief (10-15 mins) reactivation procedure (theme A for V1 & V4; theme B for V2 & V5; theme C for V3 & V6).

**Follow-up (FUP)**. Participants completed the post-intervention pain questionnaires online (from home) four weeks after the final intervention session (V6).

### Intervention Quality Monitoring

The study coordinator (ACL) completed the official Reconsolidation Therapy training course (2.5 days), and trained the other therapist (MT) accordingly. To ensure proper implementation of the protocol, an external expert in reconsolidation therapy listened to approximately 10% of the reactivation sessions (giving priority to earlier sessions) and provided feedback.

### Outcome measures

#### Feasibility

Feasibility measures were selected based on the recommendations of Thabane et al. ^45^, and include: 1) recruitment (rates and reasons for ineligibility); 2) completion of the intervention (adherence to treatment schedule, attrition, and dropouts); 3) adverse events (type, frequency, severity, attribution, expected/unexpected); 4) blinding success (perceived group allocation). Adverse events (AEs) were recorded throughout the study. For the purpose of assessing the safety of propranolol in our population, a HR lower than 55 bpm and/or a decrease of 30% in HR or BP was considered an AE. The severity of each AE was graded on a scale from 1 to 5 as per the Common Terminology Criteria for Adverse Events (CTCAE), wherein an AE of grade 1 is asymptomatic or mild, and an AE of grade 2 requires treatment ^46^.

### Acceptability

The acceptability of the intervention was assessed as per the recommendations of Sekhon et al.^47^. The five constructs assessed were: 1) burden; 2) ethics; 3) intervention consistency; 4) intervention effectiveness; and 5) self-efficacy. Each construct was assessed before and after the intervention (at V0 and FUP) with a numerical rating scale (NRS).

### Effect of the intervention

The outcome measures used to assess the effect of the intervention were selected based on the IMMPACT (Initiative on Methods, Measurement, and Pain Assessment in Clinical Trials) recommendations for clinical trials in chronic pain populations ^48,49^. The variables assessed were 1) the impact of pain on physical function, assessed using the Brief Pain Inventory – Short Form (BPI) ^50^; 2) pain severity, also assessed using the BPI; 3) emotional function, assessed using the Profile of Mood States (POMS) ^51^; 4) symptoms of central sensitization (a population-specific instrument), assessed using the CSI part A; and 5) perception of change assessed using the Patient Global Impression of Change (PGIC) ^52^. Each questionnaire was completed during the admissibility visit (V0) and at follow-up (FUP), except for the PGIC which was only completed at FUP. Additionally, the BPI was completed during each intervention session, such that partial data was available for participants lost to follow-up (however, it should be noted that reconsolidation blockade affects long-term memory only, and not short-term memory ^31^; as such, RT is not expected to decrease pain during sessions).

### Sample size

The protocol aimed to recruit 24 participants. As the primary objective of the study was to assess the feasibility of the intervention rather than obtain statistically significant results, no sample size calculation was performed. The sample size of 24 was selected based on the recommendations of Julious et al. ^53^, according to which 12 participants per group is adequate for this type of study.

### Allocation and blinding

Group allocation was performed using minimization ^54,55^, which was completed by an independent research professional using the free and open-source software MinimPy2 ^56^. The randomization index was minimal (99%) and the minimization factors were CSI score, BPI score, symptom duration, gender, and age. The participants and the therapists administering the sessions and overseeing the questionnaire completion were blinded to the group allocation.

### Statistical analysis

The feasibility indicators, such as enrollment rates and adverse events, are presented in the form of flowcharts and descriptive tables. Considering the small sample size and feasibility design, the emphasis was similarly placed on descriptive analyses for measures of acceptability and intervention effect. No interim analyses were planned or conducted.

## Results

### Sample

As planned, 24 adults with low back pain (12 per group) were recruited to participate in the study. Their average age was 42 ± 12 y.o. in the control group and 49 ± 13 y.o. in the experimental group. Both groups had seven women and five men. Additional sociodemographic information can be found in Table 2.

**Table 2.**
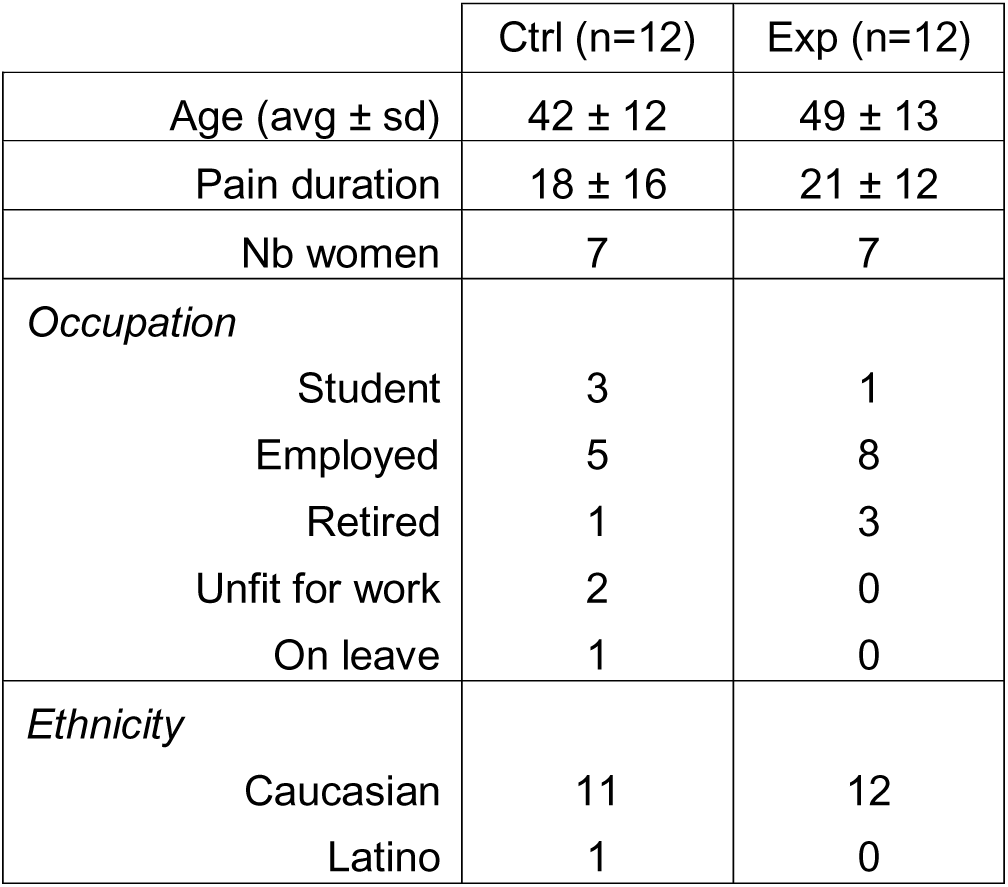
Sociodemographic Information. Table 2 shows the socio-demographic characteristics of the sample. **Abbreviations:** Exp, Experimental; Ctrl, Control

### Feasibility

#### Recruitment, adherence & retention

Sixty-six potential candidates were screened over 6 months, 35 of which completed the admissibility visit (Figure 1). Of the 24 participants enrolled in the study, 22 completed all 6 intervention visits. One participant in the control group (male, in his 30s) withdrew after the 4th visit because of emotional discomfort during the reactivation sessions, but completed the follow-up. Another participant (female, in her 60s) was excluded after the 4th visit because she started a new treatment at a pain clinic; her results related to feasibility and acceptability were analyzed as planned, but her pain scores at follow-up were not considered (analyses were conducted using her initial scores).

**Fig 1.**
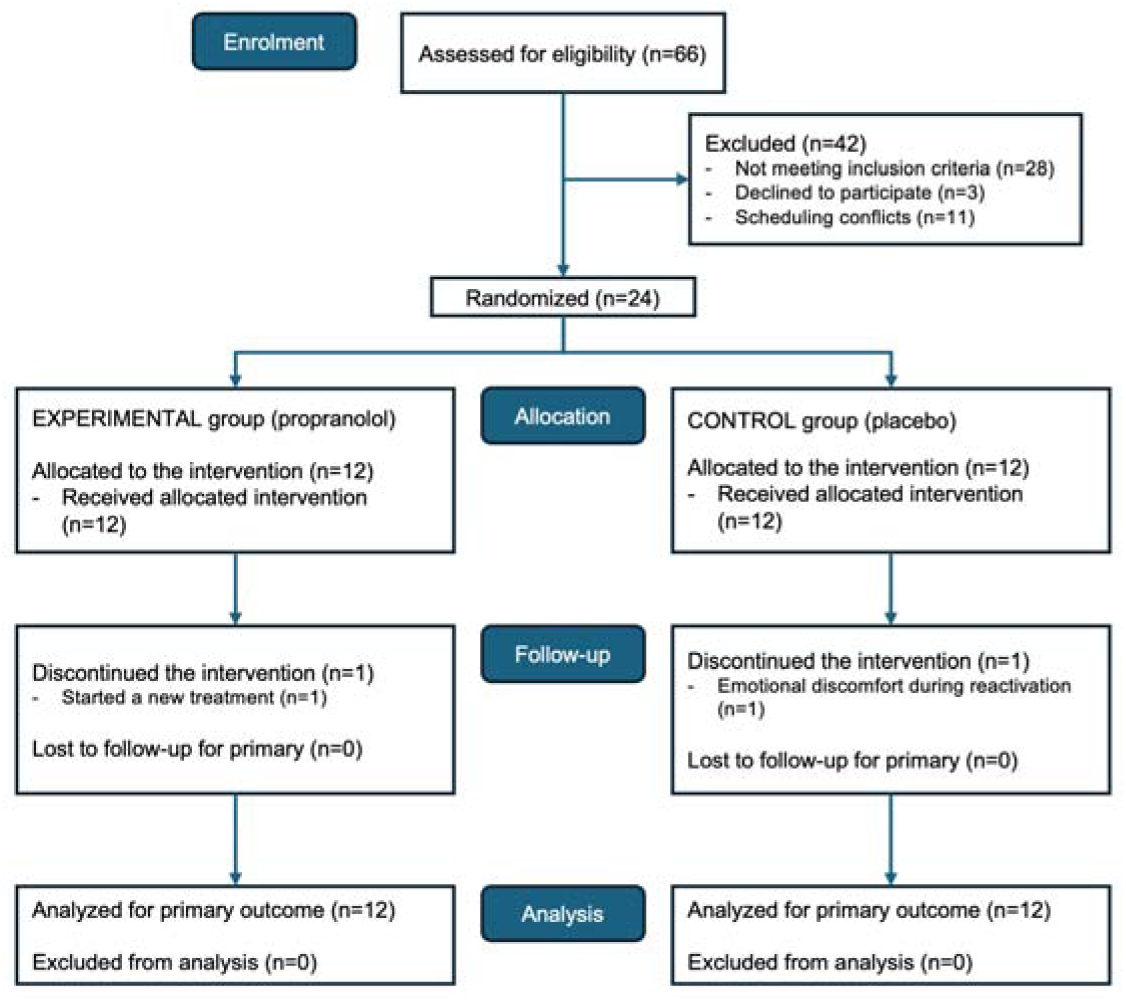
Recruitment flowchart. Fig 1 shows the recruitment flowchart for the study.

Of the 22 participants who completed all six intervention visits, 13 completed them in the planned 6-week period; 7 participants postponed one session by a week; and 2 participants postponed two sessions so the intervention took place over 8 weeks. Most postponed sessions were due to illness or holidays.

In the experimental group, 3 participants believed they received the propranolol and 5 believed they received the placebo. In the control group, 1 participant believed they received the propranolol and 6 believed they received the placebo. The remaining participants reported no suspicion either way.

### Safety & adverse events

#### Effect of propranolol on HR and BP

The variations in heart rate and blood pressure from baseline to 60 minutes after medication are summarized in Figure 2. Heart rate decreased by an average of 11 bpm in the experimental group, compared to 3 bpm in the control group (asymptomatic in all participants). No clinically meaningful changes in blood pressure were observed in either group (average change <4 mmHg).

**Fig 2.**
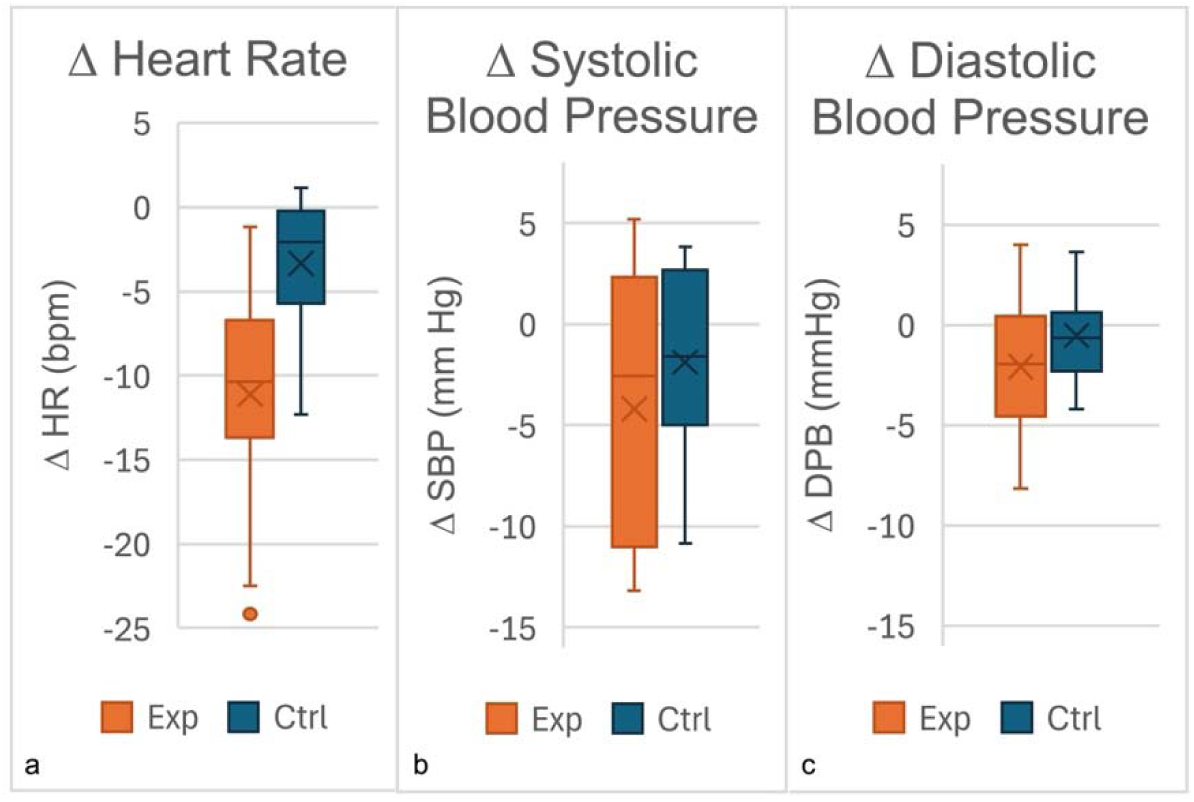
Variations in heart rate and blood pressure. Fig 2 shows the average change in heart rate (a) and blood pressure (systolic (b); diastolic (c)) from baseline to 60 mins post-medication. **Abbreviations:** Exp, Experimental; Ctrl, Control; HR, Heart rate; bpm, Beats per minute; SBP systolic blood pressure; DBP, diastolic blood pressure

#### Adverse events

AEs classified as related or possibly related to the intervention are listed in Table 3. In the control group, one participant experienced nightmares following sessions 3 and 4. In the experimental group, four participants experienced a significant (but asymptomatic) decrease in heart rate, and one participant experienced fatigue, nausea, and a headache in the evening following her second visit, but self-managed with acetaminophen and had no symptoms after her other visits.

**Table 3.** Adverse events. Adverse events observed throughout the study. Severity is classified as per the Common Terminology Criteria for Adverse Events (CTCAE). **Abbreviations**: Exp, Experimental; Ctrl, Control; HR, Heart rate; bpm, Beats per minute

|  | Description | Group | Severity (1-5) | Expected/unexpected | Session |
| --- | --- | --- | --- | --- | --- |
| L.008 | Nightmares | Ctrl | 1 | Unexpected | V3, V4 |
| L.007 | Headache, nausea, fatigue | Exp | 1 | Unexpected | V2 |
| L.059 | ↓ HR - asymptomatic (88 to 61 bpm) | Exp | 1 | Expected | V1 |
| L.048 | ↓ HR - asymptomatic (106 to 67 bpm) | Exp | 1 | Expected | V6 |
| L.019 | ↓ HR - asymptomatic (78 to 53 bpm; 66 to 52 bpm) | Exp | 1 | Expected | V5, V6 |
| L.038 | ↓ HR - asymptomatic (105 to 71 bpm; 108 to 71 bpm; 110 to 75 bpm) | Exp | 1 | Expected | V1, V2, V5 |

#### Acceptability

Five aspects of acceptability (burden, ethicality, self-efficacy, coherence, and perceived efficacy) were assessed before and after the intervention using 11-pts NRSs (0 = strongly disagree; 10 = strongly agree). The average scores for both groups were generally favorable (see Figure 3), although for both groups, the burden, coherence and perceived efficacy of the intervention tended to be rated less favorably at follow-up.

**Figure 3.**
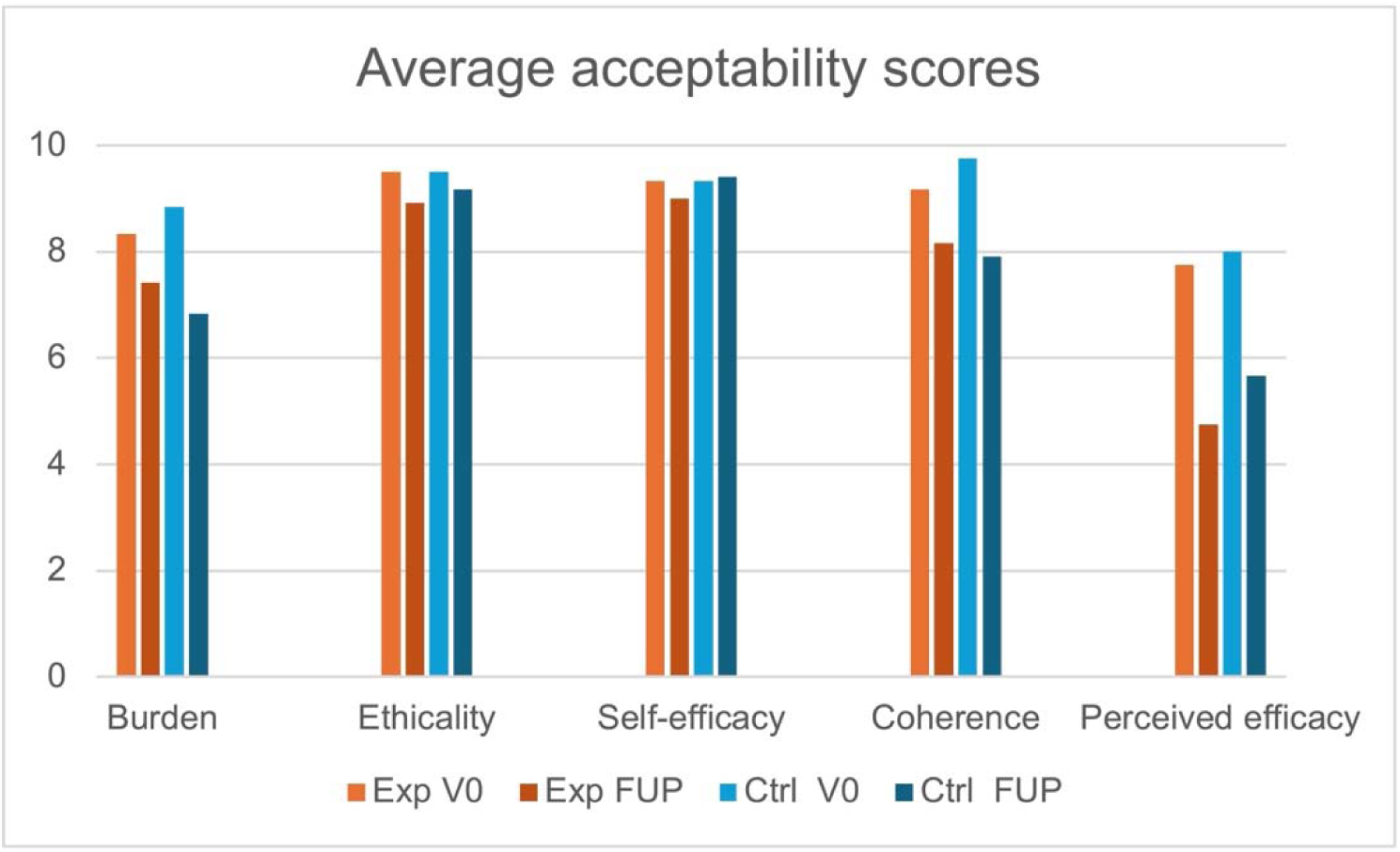
Acceptability scores. Fig 3 shows the average acceptability ratings for each of the five constructs. Ratings were obtained before the intervention (V0) and at follow-up (FUP – 4 weeks post-intervention), for both groups. Abbreviations: Exp, Experimental; Ctrl, Control; FUP, follow-up

#### Effect of the intervention

The effect of treatment on symptoms was evaluated using the BPI-intensity, BPI-function, POMS, CSI, and PGIC. Figure 4 shows the change in individual scores for each questionnaire, with negative values indicating a decrease in symptoms (ie, an improvement). Approximately half of the participants in both groups experienced a clinically significant improvement in physical function (≥ 1/10); other outcomes remained stable on average and were similar across the two groups, suggesting no superior effect of the experimental intervention over the placebo intervention (Fig 4).

**Fig 4.**
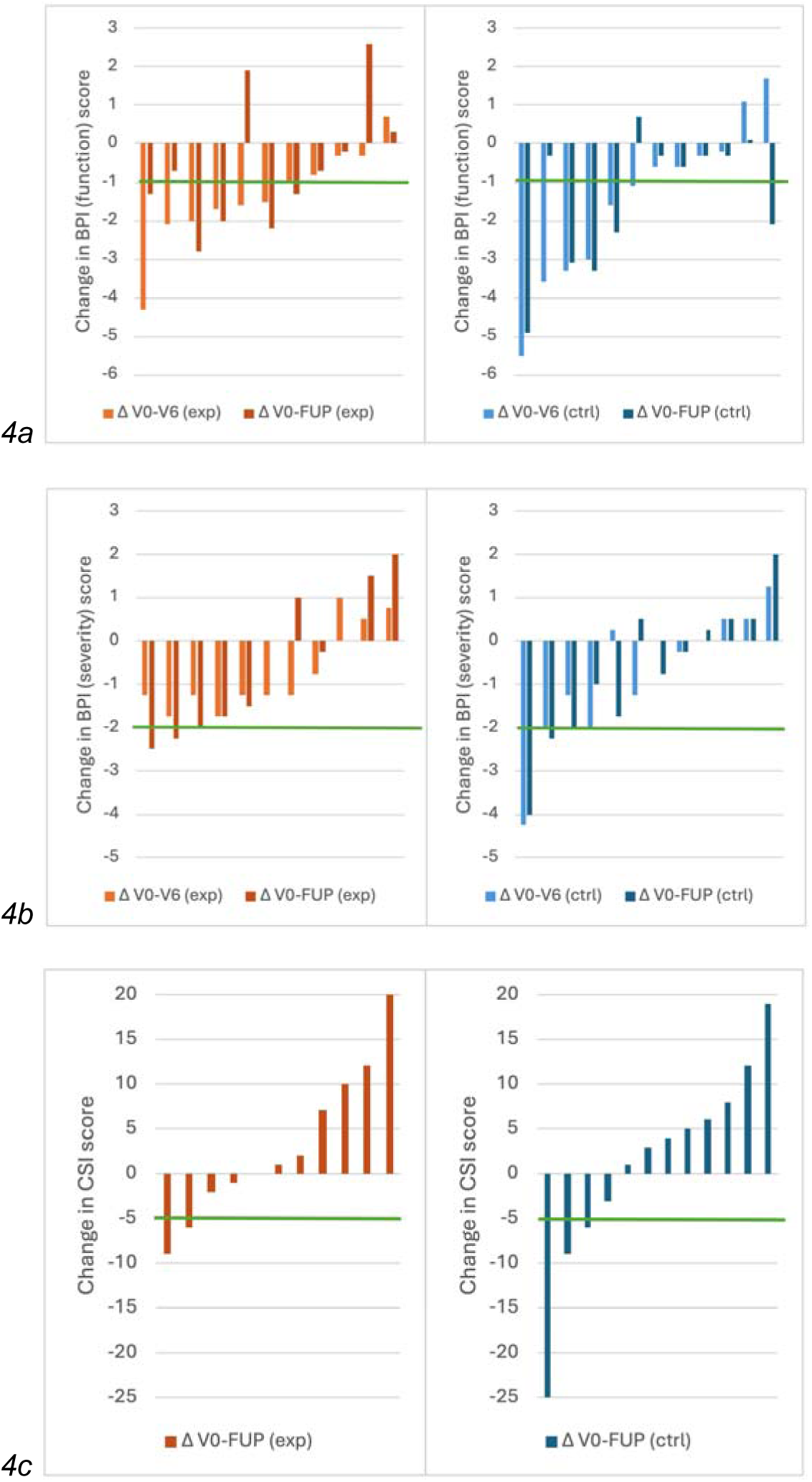
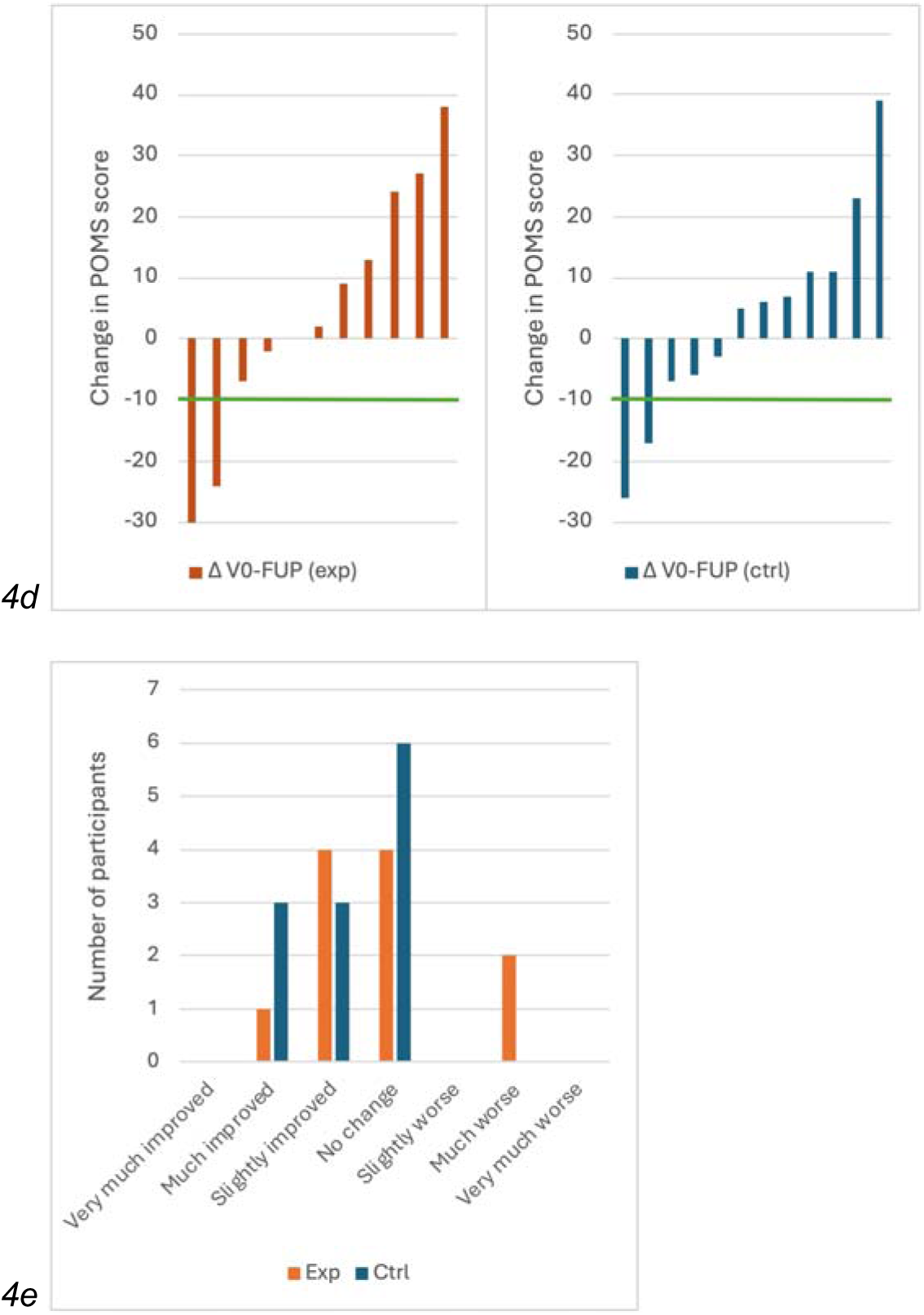
Effect of the intervention. Fig 4 shows the individual changes observed in each participant for each outcome measure (Brief Pain Inventory – Function (a); Brief Pain Inventory – Intensity (b); Central Sensitization Inventory (c); Profile of Mood States (d); Patient Global Impression of Change (e). Negative scores represent reductions in symptoms, ie, improvements (Δ = FUP – V0). The green line indicates the threshold for clinical significance for each instrument. **Abbreviations:** Exp, Experimental; Ctrl, Control; FUP, follow-up; Fct, function; Int, Intensity; CSI, Central Sensitization Inventory; POMS, Profile of Mood States; PGIC, Patient Global Impression of Change

#### Incidental findings

Two unforeseen findings related to the adequacy of the intervention emerged during the study. The first concerned participants’ understanding of their pain condition, and the second their response to the reactivation procedures.

#### Pain conceptualization

No formal measure was used to assess participant’s understanding of the mechanism underlying their low back pain. However, throughout the study, the language used by participants to describe their condition suggested that they conceptualized their pain as stemming from a musculoskeletal lesion, and as being an adequate indicator of tissue damage. For instance, most participants described their pain using language suggesting a strong belief that their pain originated from a musculoskeletal lesion – citing elements such as osteoarthritis, a ‘weak spine’, displaced vertebrae, etc. Such explicit beliefs about tissue damage and risk of injury may have been a major confounding factor in this study, as discussed below.

### Reactivation procedures

Different issues were also noted with each of the 3 reactivation “themes”.

#### Theme A (worst pain episode)

This theme, wherein participants were asked to describe their most painful episode, followed the PTSD protocol as closely as possible – including instructions to describe the environment and all sensory experiences associated with the event. Participants could easily identify and describe their most painful event, they often appeared unsettled when asked to describe the environment, the sounds and smells associated with the event. Most participants reported that these prompts were jarring; not only could they seldom remember those details, having to focus on these (perceived) irrelevant details also ‘pulled them out’ of the immersion.

#### Theme B (“worst nightmare”)

For this theme participants had to make up and picture a ‘nightmare scenario’ related to their pain – in other words, to picture doing an activity they expected would cause pain. However, several participants unexpectedly described a scenario where pain ended up not being the primary concern (eg, waking up on a particularly bad day, lying in bed unable to move because of excruciating, debilitating pain – then hearing their child being struck by a car and being unable to provide assistance because of the severity of their pain). For others, the implausibility of performing or carrying on with a pain-inducing activity (instead of resting, stopping, or modifying the activity) appeared to prevent full immersion.

#### Theme C (negative emotions)

This theme aimed to reactivate negative emotions associated with pain. Based on our patient-partner pre-tests, we expected that patients would describe emotions such as distress, anxiety, and pain-related fear. However, the majority of participants spontaneously described emotions related to the consequences of the pain (eg, guilt over not being able to fulfill their role as parents or spouses; fear that they might end up in a wheelchair; frustration with the healthcare system), rather than related to the pain itself.

## Discussion

The objective of this study was to assess the feasibility and acceptability of reconsolidation therapy as a treatment for chronic nociplastic low back pain, and to gather preliminary data on the effect of the intervention on patient-reported pain symptoms.

### Feasibility

The intervention appears safe, with only mild and infrequent adverse events in the experimental group. These results are unsurprising given propranolol’s favorable safety profile ^44^; however, seeing as the medication was used off-label, and for the first time with people suffering from low back pain, documenting its safety was crucial. There were relatively few dropouts (n = 1 in each group), and the participants generally adhered to the treatment schedule: the majority of participants completed their 6 visits over the planned 6 weeks, and only 2 participants required the maximum allowed time of 8 weeks to complete their 6 visits. These results suggest that the intervention is feasible with this population.

### Acceptability

The results regarding acceptability were favorable, with the notable exception of perceived efficacy, which was rated lower by both groups following the intervention. This may have been due to the lack of improvement experienced by most participants. However, these generally high acceptability scores should be regarded cautiously, as they were likely inflated by a sampling bias – the prospective participants who judged the study unacceptable after seeing the recruitment posters presumably chose not to contact our research team, such that their unfavorable opinions remain undocumented. Moreover, acceptability measures have advanced considerably since the experimental protocol was developed. The design of future acceptability studies should take advantage of these new tools.

### Pain education

Some of the improvements observed in certain participants may have been due to pain education. However, the absence of clinically significant improvement in the control group – and, indirectly, the persistence of beliefs equating pain to tissue damage, despite educational content on nociplastic pain – suggests that the videos did not have a clinically meaningful impact, on average. Future studies could allocate additional resources to pain education, for example by adding prompts and quizzes throughout the video (to improve engagement and monitor adherence), or by having the education delivered in person by a healthcare professional.

### Effect of the intervention

Our findings suggest that, in this particular form and application, RT had no impact on pain symptoms, as we did not observe a clinically meaningful pattern suggesting that the experimental group experienced a greater reduction in symptoms compared to the control group. However, this does not constitute robust proof that RT cannot be an effective treatment for chronic nociplastic pain. Indeed, as discussed above, the activation procedures used may have failed to fully reactivate the target synapses – which would mean there was no reconsolidation for propranolol to interfere with. It is also possible that the success of RT depends on participants not holding a strong belief that their pain indicates tissue damage – a prerequisite that was not met in the present study. Therefore, it is still possible that another version of RT, properly accounting for these two potential confounding factors, could benefit patients suffering from chronic pain (see below).

### Incidental findings – adequacy of the intervention

#### Pain beliefs & conceptualization

The language used by participants suggests that they conceptualized their pain as arising from (and signalling) tissue damage to their lumbar spine. In other words, they appeared to hold *explicit beliefs* that their spine’s physical integrity was compromised. This could represent a fundamental (and not previously considered) difference between patients with chronic low-back pain and those with PTSD, the latter likely being more aware that their fear reactions are excessive and disproportionate to the actual threat level. The explicit belief that pain stems from physical injury could have affected the efficacy of the intervention. Indeed, reconsolidation blockade with propranolol does not target (explicit) declarative or episodic memory, but rather targets (implicit) associations encoded in the amygdala ^57,58^. Moreover, research on the nocebo effect has shown that the mere belief that something is harmful or causes pain can lead to pain, even in the absence of actual damage or threat of damage ^59–61^. As such, holding a belief that pain reflects physical injury could be sufficient to trigger and maintain the chronic nociplastic pain state – regardless of any possible weakening effect of reconsolidation blockade on implicit associations in the amygdala. In other words, it is possible that reconsolidation blockade can only benefit patients who recognize that their pain is maladaptive and arises from a dysregulation in the nervous system – a prerequisite that was unmet in the present study. To clarify this matter, future studies are needed that either provide more pain education, have inclusion criteria related to pain beliefs, or formally assess pain beliefs to determine whether they are indeed a prerequisite for RT success.

#### Reactivation procedures

As outlined in the results, despite the adaptations implemented, the reactivation protocols employed in the present study appear to have been suboptimal for our population. This could have hindered the effect of the intervention, as reactivation is a fundamental aspect of RT. Indeed, only synapses that have been specifically reactivated return to a labile state and are susceptible to reconsolidation blockade, and a number of (often subtle) boundary conditions must be met for reactivation to be effective ^62–64^. For instance, while the feelings of guilt and helplessness evoked in many participants during Theme C are valid and understandable, they may not be as closely linked to the amygdala as fear of injury or pain (the intended target for Theme C). However, now that the shortcomings of the current reactivation procedures have been documented, future studies should attempt to further depart from the PTSD protocol. Specifically, they should modify the reactivation procedures to tailor them more specifically to the pain population, for example by focusing on daily painful experiences or mental imagery of painful activities.

#### Additional considerations for future studies

Despite the plausibility of its mechanism of action, RT remains relatively unknown outside of psychotraumatology and is mostly absent from the pain literature (with the notable exception of an elegant pre-clinical study by Bonin and De Koninck targeting spinal nociceptive circuits ^65^). Now that the general feasibility of RT has been established and that the safety of off-label propranolol in patients with chronic pain has been documented, a next step might be to conduct imaging studies to confirm the mechanistic plausibility of the intervention. Indeed, the intervention postulates that reactivation procedures activate the amygdala (necessary to trigger synaptic reconsolidation), but this has never been validated and will need to be confirmed. Such mechanistic studies should also assess different reactivation methods (verbal descriptions *vs* visual imagery; focusing on specific pain episodes *vs* typically painful activities, etc.) to identify those that most reliably activate the amygdala in patients with chronic pain. The mechanism of action of RT also postulates that propranolol acts specifically by blocking reconsolidation in the amygdala. This assumption is largely based on preclinical studies showing similar effects whether propranolol is administered systemically ^27^ or directly into the amygdala ^31^, with similar outcomes observed using other reconsolidation-blocking agent such as anisomycine ^26^. However, this mechanism will need to be validated in pain as well.

The present study was designed before criteria for nociplastic pain were established criteria ^66^, and future studies should obviously follow the latest recommendations when establishing their selection criteria. RT might also be studied in other populations of nociplastic pain, such as women with vestibulodynia or patients with comorbid nociplastic pain and PTSD.

## Conclusion

This was a pioneering study – the first, to our knowledge, to study reconsolidation therapy as a possible treatment for chronic pain. It provides the very first data regarding the use of this intervention in that population. Notably, our results support the safety of the intervention, an important first step in any investigation involving the off-label use of medication. Moreover, while results relating to the effect of the intervention failed to show any benefit of RT in this specific form, two major possible confounding factors were identified – both of which can be addressed by reworking the intervention protocol. As such, our findings provide a strong foundation for future studies, reflecting the iterative nature of research and highlights the importance of running pilot studies before embarking in large randomized trials.

## Data Availability

All data produced in the present study are available upon reasonable request to the authors

## Acknowledgments

The authors would like to thank Virginie Carrier, the study monitor, for her outstanding competence and continued support. They would also like to thank Marilyn Tousignant for her role as therapist; Michelle Lonergan for her role as reconsolidation expert; Jonathan Landry for his contribution to the production of the pain education videos; and Mélanie Morin and Martine Bordeleau for their technical advising during study design.

## Disclosure

The first author is supported by a Fellowship from the Canadian Institutes of Health Research Fellowship; the study was funded by the Quebec Pain Research Network (Fonds de recherche du Québec – Santé); and GL received salary support by the Fonds de recherche du Québec – Santé (Senior Clinical Research Scholar).

AB teaches reconsolidation therapy for the treatment of PTSD. There are no other conflicts of interest to declare.

The data used in the present study are available from the authors upon request.

